# Transdisciplinary Epidemiology in Schools: Integrating Molecular Environmental and Social Surveillance Through Community Science

**DOI:** 10.64898/2026.05.06.26352508

**Authors:** Orsolya Molnar, Anna Schedl, Donata Giulini, Gergely Odor, Tobias Fragner, Matthew Thornton, Karin Garber-Pawlik, Brigitte Gschmeidler, Silvia Prieler, Bettina Girschick, Veronika Kunnert, Brigitte Schmidt, Daniel Hackl, Anna Golos, Fanny Scharf, Nils Bach, Hanna Raith, Anika Stuebegger, Matthias Trenker, Janet Schiefer, Vanessa Dascalescu, Wead Asu Osaid, Christina Margioulas, Christine Marizzi, Fabian Amman, Igor Grabovac, Andreas Bergthaler

## Abstract

Respiratory virus surveillance is often constrained by symptomatic testing and centralized sampling, producing blind spots in populations at risk. We developed a transdisciplinary community science framework in which students and teachers co-designed and implemented decentralized respiratory surveillance integrating environmental, molecular, and social data in schools. Within this participatory setting, indoor air COLJ concentrations were monitored alongside student-collected air filter and surface samples, analyzed by digital PCR and sequencing. Community-generated samples reliably captured circulating viruses, while COLJ measurements revealed associations between indoor air quality and pathogen abundance. Quantitative surveys identified social and structural barriers to preventive measures, and shared ownership of study design fostered sustained engagement. By embedding epidemiology within a co-developed, community-driven research process, this study demonstrates how scalable surveillance can close critical data gaps. Our work provides a blueprint for decentralized, community-engaged infectious disease monitoring that positions local partners as active contributors to public health intelligence.

## Introduction

Seasonal respiratory viral diseases pose a significant burden on healthcare systems and broader economic development. Their potential for producing global pandemics makes them one of the most serious public health threats, raising international concern^1,2^. Given that transmission dynamics are shaped by genetic, socio-economic, and environmental characteristics,^3,4^ recent calls have highlighted the crucial role of early detection. However, traditional surveillance methods often cannot keep up with complex transmission dynamics in the absence of comprehensive databases that include molecular, environmental, and societal drivers ^5–7^. Epidemiological surveillance, therefore, needs new and cost-effective approaches to address data gaps.

Traditionally, molecular epidemiological data is patient data originating from symptomatic individuals seeking healthcare services, relying on both access to services and symptom-driven healthcare-seeking behavior.^8,9^ Environmental surveillance offers an opportunity to circumvent such barriers and collect specimens from a more representative population sample.^10^ Wastewater-based epidemiology (WBE), a central tool for viral surveillance during the Coronavirus Disease 2019 (COVID-19) pandemic, offers powerful and robust alternatives for population-level surveillance but has limitations, including inflexible granularity, particularly in underdeveloped areas, and lack of non-geographically stratified data of disproportionately affected subpopulations^11–13^. Air filtering devices enable collecting data directly from spaces frequented by high-risk communities in a non-invasive manner. Thus, to gain crucial insights into respiratory viruses circulating in high-risk groups, air sampling has recently been adopted to survey schools, hospital wards, and student dorms^14,15^. Airborne pathogen load has been hypothesized to covary with other measures of air quality, particularly levels of CO ^11,16,17^. High indoor CO_2_ levels in turn cause physiological and neurological symptoms due to increasing rates of rebreathing exhaled air or “shared air”^18^. Indoor air ventilation has therefore gained attention as an important prevention measure complementing primary prevention in confined spaces such as hospitals and schools^19,20^. Using air filters and CO_2_ sensors, indoor spaces can provide valuable insights into the respiratory pathogen burden and air quality in public and community spaces, such as school settings.

However, the absence of a social dimension in molecular datasets impedes the development of targeted programs that empower communities facing higher infection risk^21,22^. Public educational settings elevate the risk of respiratory infections for both children and teaching staff, building better environments (hospitals, schools, etc.) is therefore one of the key priorities of pandemic preparedeness^23,24^. Among the multitude of groups underrepresented in traditional surveillance methods, school communities are of particular interest from both scientific and public health perspectives. They spend extended periods in confined spaces, creating highly interconnected transmission networks that foster potential outbreaks^25^ and transferring them to more susceptible age groups (e.g., <5 or >55 yrs) within their residential community^26^. Adolescents may also exhibit lower compliance with infection prevention and lower health literacy^27,28^. Utilizing the structured environments of schools, students are specifically targeted by primary prevention programs and awareness-raising campaigns about prevention measures^29,30^, but their perception of and adherence to prevention measures are determining factors in the success of managing outbreaks.

To deliver both data and impact, novel epidemiological studies need to partner *with*, and not go in and extract information from communities to benefit both science and society. Transdisciplinary frameworks help design participatory initiatives in which stakeholders co-design studies at eye level^31,32^. Community Science (CS) actively includes the general public in the research process. By empowering communities to co-create and benefit from information, this approach increases access to science and promotes empowerment, trust, and research that addresses social needs. However, other terminologies like participatory science or citizen science are also legitimate and might be more suitable in some contexts^33,34^. CS programs assign expert roles to community members within a scientific context, enabling researchers and laypeople to jointly address disparities, the underrepresentation of marginalized groups, and implementation barriers^35^. The risk group of adolescents often faces a lack of ownership and agency in their health decisions, which forms barriers to mitigating infections^36,37^. Community science empowers communities by incorporating lived experience, community values, and local conditions for long-term impact^35,38,39^. Empowering students through participatory initiatives helps prevent infection, yields epidemiological data and improves health outcomes,^40,41^ potentially advancing Goal #3 (Good Health and Wellbeing) and Goal #11 (Sustainable Cities and Communities) of the United Nations Sustainable Development Goals^35,38^.

Co-developmental or extreme citizen science is globally underutilized in epidemiological surveillance despite its benefits, such as filling molecular data gaps and improving preventive measures through higher compliance rates and better policy implementation^40^. Our study delivers a much-needed proof-of-concept that a comprehensive Community Science program can yield targeted environmental, molecular, and socio-epidemiological data through transdisciplinary engagement with stakeholders, thereby acquiring samples unavailable through extractive disease surveillance.

We developed a community science program, LuPFiS (“**Lu**ftpartikel und **P**athogen **Fi**lteruntersuchung mit **S**chüler*innen”; “Air Particle and Pathogen Filter Investigation by Students”), to position high school student communities as active contributors to integrated environmental, molecular, and social data collection within their own schools. By uniting environmental exposure metrics, molecular pathogen detection, and social and behavioural context data collected directly within school communities, the approach enables an unusually high-resolution, interdisciplinary understanding of transmission risks that cannot be captured by single-domain methods. This integration at the community level provides a more holistic depiction of infectious disease determinants, illuminating interactions between air quality, pathogen presence, and local practices. LuPFiS integrates environmental, molecular, and socio-epidemiological analyses into an innovative, inter- and transdisciplinary framework to concurrently generate comprehensive epidemiological datasets and foster agency and empowerment in their health outcomes among students.

Our program aimed to test whether data collected by non-professional scientists shows sufficient consistency and robustness to be integrated and statistically compared with traditional surveillance data, leveraging the benefits of comprehensive datasets provided by communities. We designed high-quality data collection pipelines to acquire CO_2_ measurements, pathogen samples, and social questionnaires, simultaneously obtaining information on air quality, circulating respiratory virus abundance and diversity, and implementation barriers to prevention measures. To build a reproducible and scalable model for inclusive and replicable epidemiological monitoring, we developed our program to focus on generating scientific knowledge while empowering marginalized voices^42^, while documenting key pillars and milestones of a transdisciplinary framework enriched by quantitative tracking of participant recruitment and engagement^43^.

## Results

We established a unified pipeline that integrates environmental, molecular, and socio-epidemiological data streams into a consistent framework for school-based infectious disease surveillance (Fig. 1). The process spans recruitment and co-development through implementation and dissemination, with researchers, teachers, and students jointly involved at each stage. Environmental surveillance generated CO_2_-derived indicators of exposure risk, molecular analyses produced quantitative and compositional measures of viral burden, and social sampling captured behavioral and perceptual correlates of prevention. Standardized laboratory and analytical procedures ensured that each data stream produced harmonized readouts—viral abundance and richness, air-quality metrics, and social response indices—that could be jointly interpreted. This integrated design provides a reproducible template for linking physical, biological, and social determinants of infection within a single transdisciplinary surveillance framework.

**Figure 1.**
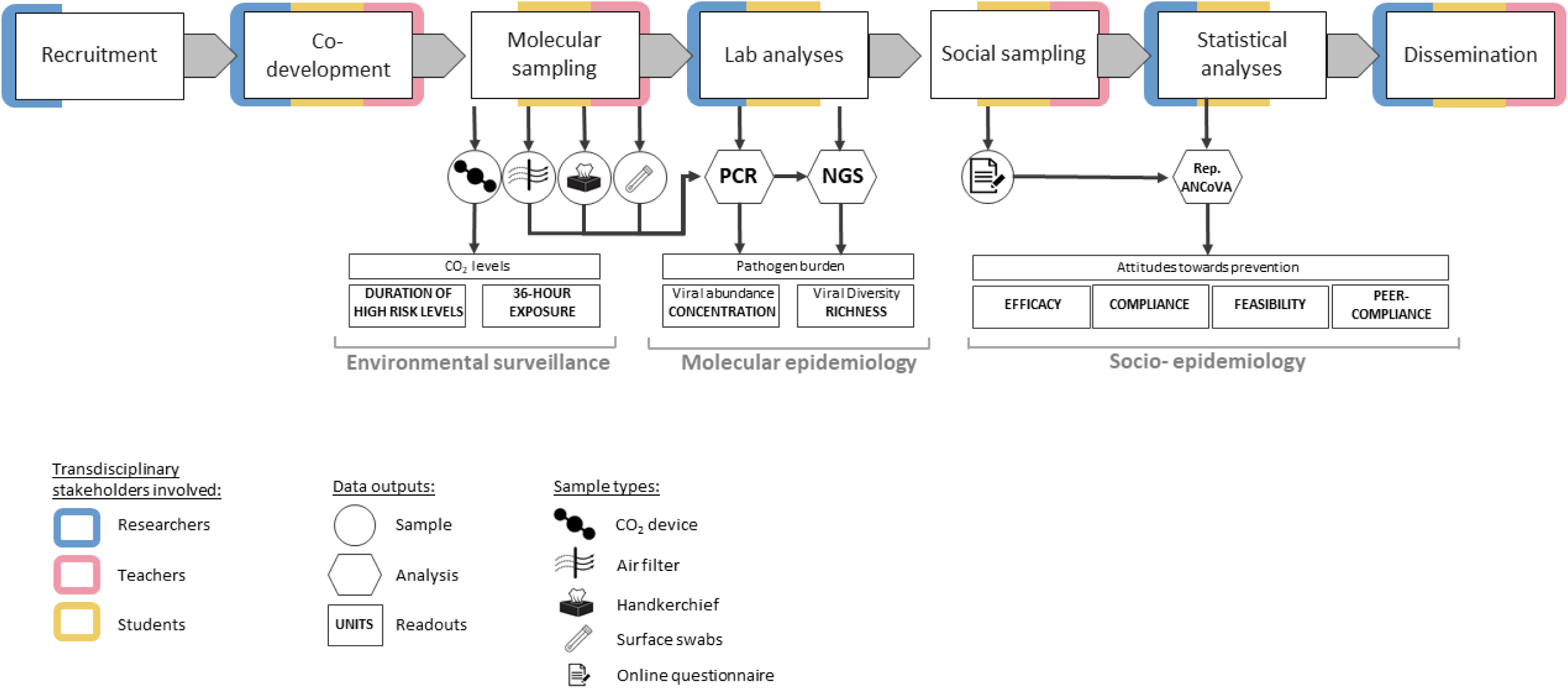
**Workflow of the LuPFiS study**. Stages of the LuPFiS community science programme are shown in boxes, with frame colours indicating the stakeholder groups involved in design or execution. Data outputs are marked by circles, with icons denoting sample type. Analyses performed on each output are shown in hexagons, and resulting readouts in rectangles, with measurement units given in bold capitals.

### Environment affects pathogen abundance more than diversity

Molecular sampling tested air filter, surface swab, and disposable handkerchief samples to characterize the abundance and richness of respiratory pathogens in indoor school spaces. When estimating pathogen abundance, disposable handkerchief samples yielded inconsistent sampling frequency (Supplementary Figure 1) as well as several-fold higher copy numbers (Supplementary Figure 2A).

To characterize airborne pathogen burden, the relative abundance of PCR targets was calculated for each date as the sum of viral copy numbers from air-filter and surface-swab samples for each school. When analyzed against a time scale, we detected a temporal shift from RSV being the most abundant pathogen in Phase 1 schools to SARS-CoV-2 being more frequent in Phase 2 schools (Fig. 2B).

**Figure 2.**
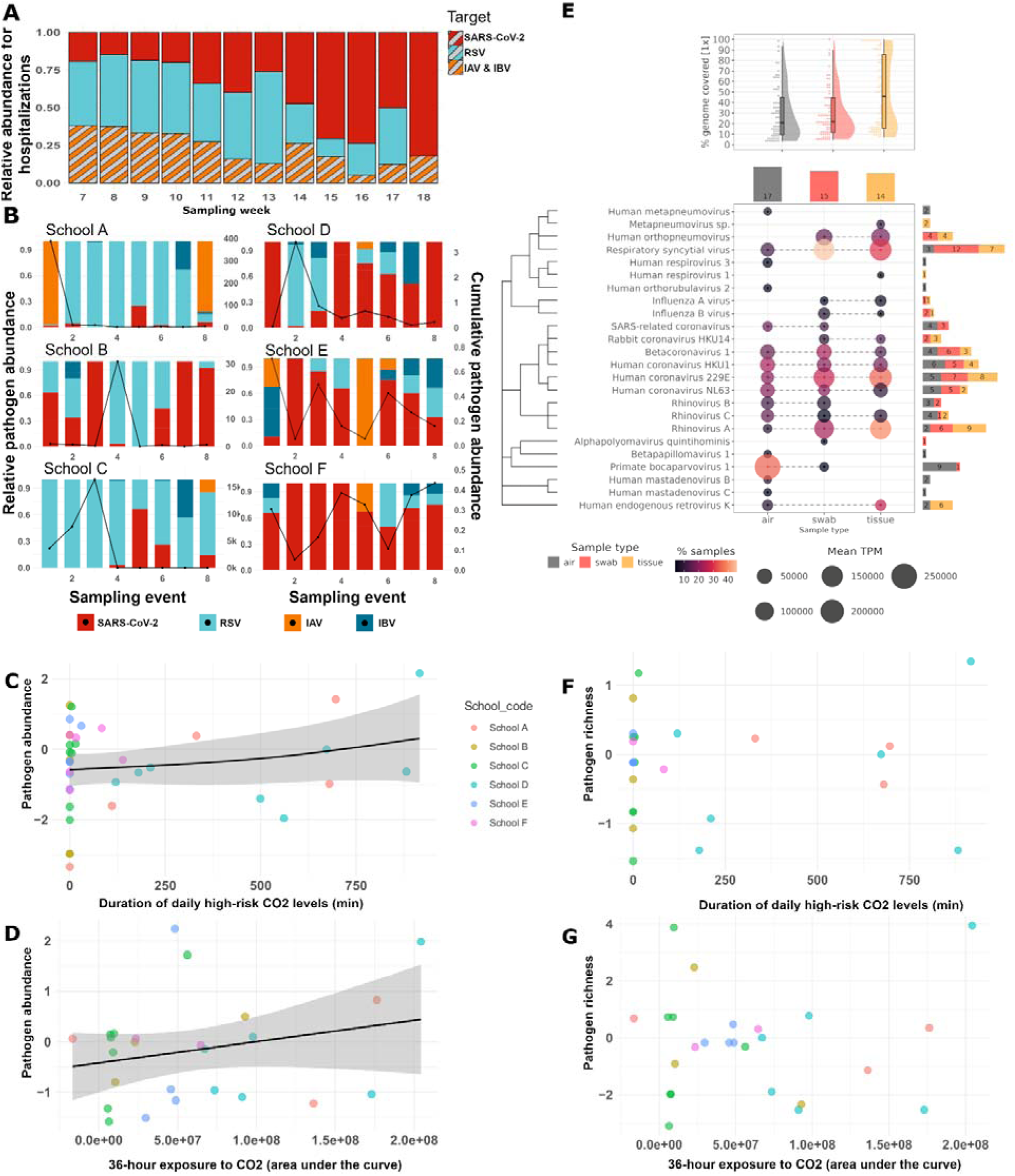
**Environmental factors associated with pathogen abundance and diversity**. Hospitalization rates in Vienna for SARS-CoV-2, RSV and Influenza A and B viruses (IAV&IBV) combined (A) are shown alongside PCR-derived abundances of SARS-CoV-2, RSV, IAV and IAB across six schools (B). Hospitalization data are aligned to school sampling weeks for Phase 1 (B left) and Phase 2 (B right). Line graphs overlay relative abundance plots to indicate cumulative pathogen load per sampling event. Genome coverages (% of genome with ≥1 read) are summarized per sample type in the violin plots with each dot corresponding to a single reference sequence. Per species metrics are summarized in the bubble plot with bubble size indicating the mean TPM value and color denoting the proportion of samples detecting the species for the respective sample type. Raw detection counts are shown in the right-margin barplot and total sample numbers per sample type in the top-margin barplot. Viral abundance was positively associated with both duration of high-risk COLJ exposure (C) and 36-h cumulative COLJ exposure (D), whereas neither metric correlated with viral richness (F, G).

Patient data from Austrian hospitals showed comparable temporal trends (Fig. 2A) ^44^, demonstrating that student-collected samples can be analyzed within the same pipeline as conventional surveillance data. This comparability enables direct evaluation of how school environments contribute to regional pathogen dynamics.

Pathogen diversity was estimated through sequencing of all three sample types. The number of targets detected in each sample contributed to viral richness, which was used to characterize diversity. Due to limited availability of handkerchief samples, sample type had no detectable effect on pathogen diversity, which led to all sample types being entered into future analyses (Supplementary Figure 2B). We detected 24 viral species across all sample types. Common respiratory pathogens such as Respiratory Syncytial Virus (RSV), human coronavirus 229E, and rhinoviruses were abundant across locations and sample types, while the unexpected detection of rabbit coronavirus HKU14 highlights the utility of sequencing as a multiplex detection tool in school-based surveillance (Figure 2E). Since viral richness was not affected by sample type, diversity analyses can benefit from a wide range of sampling methods,

Indoor air quality was described by the duration of high-risk CO_2_ levels (Duration) and total exposure (Exposure) over 36 hours. While Duration shows the time during which CO_2_ levels pose a health risk, Exposure shows level of CO_2_ to which students and staff are exposed. Duration (Fig. 2C, Supplementary Figure 3A) and Exposure (Fig. 2D, Supplementary Figure 3B) showed a significant, positive association with viral abundance, but not with richness (Fig. 2G-H, Supplementary Figure 3C-D). CO_2_ levels are commonly referred to as a proxy for pathogen load^16,17,45^, but understanding pathways through which they affect transmissibility, measures associated with abundance and richness need to be disentangled.

### Prevention measures face either feasibility or compliance barriers

Given that most viral pathogens detected in school settings are vaccine-preventable and that schools are often primary targets of vaccination campaigns, our surveys aimed to reveal attitudes towards primary prevention methods among students. Surveys yielded a total of 242 fully completed questionnaires collected from five different schools. Repeated-measures ANCOVA on factors influencing vaccination showed that parental advice has a stronger effect on attitude towards vaccination than peers or social norms (Supplementary Figure 4). This aligns with previous results indicating that family relationships have a strong influence on attitudes towards vaccination^46^. Vaccination was perceived as an efficient prevention method, with high levels of self-admitted compliance and feasibility, but peer compliance was perceived as lower, with no gender effect (Fig. 3A). This result should be further tested across a broader range of student populations, given the high rates of vaccine hesitancy Austria is facing^47^. Social distancing and absence during sickness were both perceived as efficient, but showed low self-admitted compliance, low feasibility, and lower peer-compliance (Fig. 3 B-C). While absences were judged similarly by all genders, social distancing showed gender-specific differences (MS = 4.730, F = 7.436, P < 0.001). Females reported higher perceived efficiency and self-compliance with lower perceived peer-compliance, while males estimated efficiency lower and self-compliance lower, and peer-compliance higher (Fig. 3 B). As females reported stronger negative impacts during COVID-19 measures ^48^, they may overestimate the effort required to maintain social distancing, resulting in higher perceived efficiency and compliance. This result requires comparing self-admitted compliance levels with the actual adherence rate to distancing measures as an independent control. Absenteeism during sickness was perceived as efficient, but showed lower compliance, lower feasibility, and lower peer-compliance, most likely because absence during short-term illness is subject to caregiver approval and is therefore not the individual choice of respondents.

**Figure 3.**
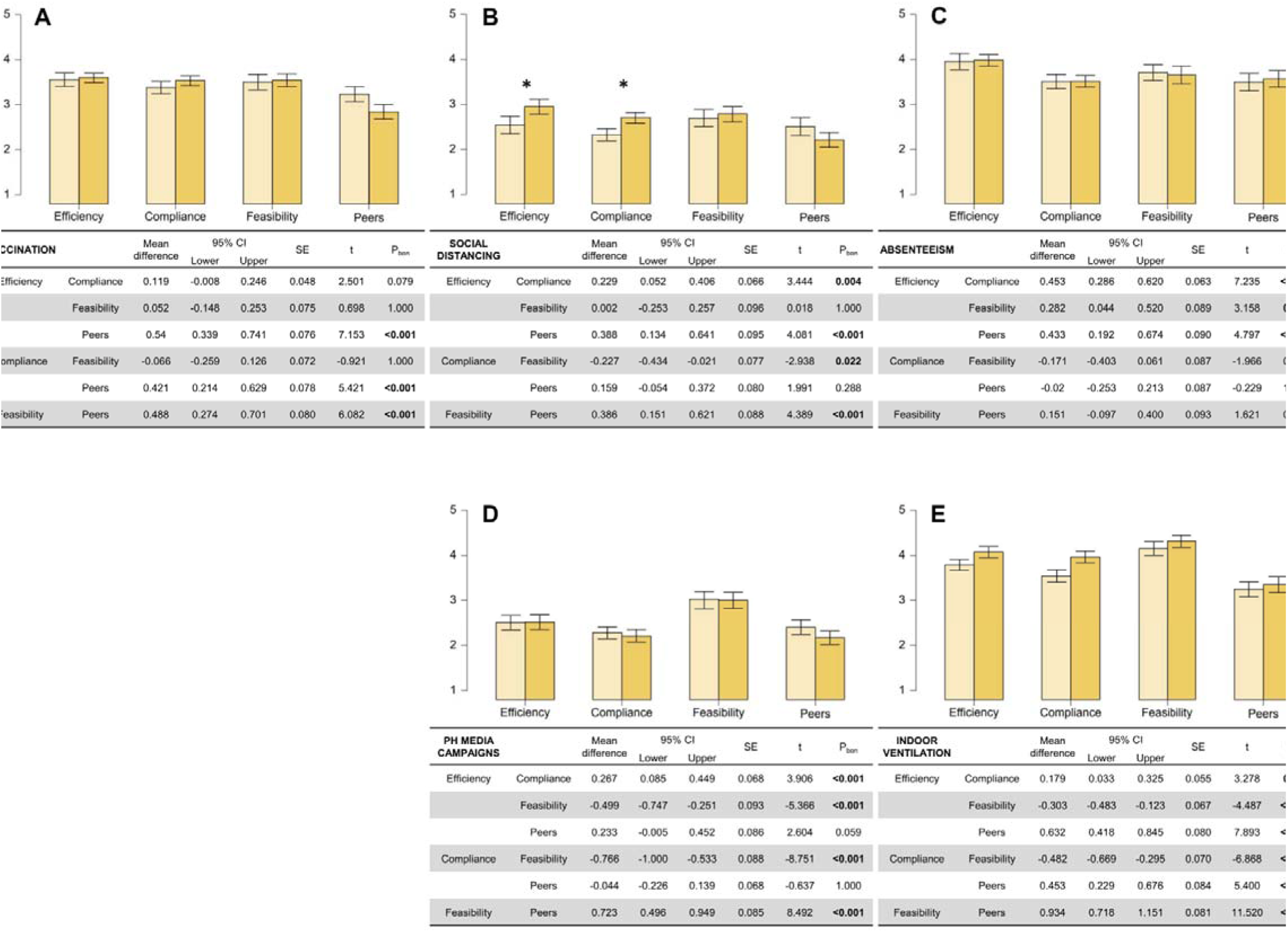
Perceived feasibility and compliance barriers to prevention measures among student stakeholder group. Vaccination was perceived as efficient, feasible and highly compliant (A). By contrast, social distancing (B) and absence during sickness (C) were judged efficient but of low feasibility and compliance. Efficiency and compliance with social-distancing was ranked higher by females (yellow bars). Public health media campaigns (D) and indoor ventilation (E) were considered feasible but inefficient, and therefore of low compliance. Effects are displayed for females (yellow bars) and males (beige bars) separately, asterisk above bars indicates significant differences between genders. Post-hoc tables display significance levels with Bonferroni corrections in bold for within subject effects (P < 0.05)

Attitudes towards public health announcements in print, broadcast and digital media and indoor ventilation showed different trends: both were perceived as highly feasible but low in perceived efficiency and in self- and peer compliance (Fig. 2D-E). Public health announcements achieved the lowest compliance score of all prevention measures tested, showing students are likely disconnected from and unaffected by regular media announcements. Similar patterns of declining responsiveness to public-health information have been documented during the pandemic,^49^ suggesting that students may likewise become less receptive to, or even negatively affected by, repeated media announcements. Although ventilation was perceived as less efficient with lower overall compliance, the overall scores were highest for this preventive measure. This could indicate that compliance is driven not by students but by teachers reminding classes to ventilate. Overall, results highlight that different preventive measures face either feasibility or compliance issues and can advise awareness-raising programs on how to improve implementation.

When asked about symptoms affecting absenteeism, fever and sore throat had a positive effect, while coughing and increased nasal discharge did not (Supplementary Figure 4).

### Flexibility and leadership roles foster continued engagement and continuity in community science programs

We sought to systematically track evolving network dynamics and pathways of knowledge exchange throughout the community science program to elucidate stakeholder interactions and quantify patterns of engagement. Network visualization showed that involving key stakeholders connected to educational experts allows for synchronized and broad recruitment of teachers, which in turn facilitates student enrollment (Fig. 4A – network 1). Engagement tracking showed that different phases of the study saw different participants from the same stakeholder group assume a more dominant role, with different students and teachers being more active in different tasks (Fig. 4A – network 2-4). By allowing flexible commitment across the program’s different phases, participants can stay motivated and adapt their engagement routine and schedule to shorter, but deeper, engagement periods. Survey respondents mapped against survey developers’ points to the efficiency of community and citizen science networks, where five active student developers reached 242 respondents (Fig. 4A – network 5). An established stakeholder network can amplify efforts to gather community-specific data and assist policy implementation by incorporating lived experiences and fairness^35,38^. Network 6 on Fig. 4A shows that disseminating results back to schools led to incoming students attending the workshops, ensuring continuity between classes of different years. School-based participatory science programs can foster engagement and continuity by disseminating results back, to provide an opportunity for new participants to replace outgoing classes. The participation rates of different stakeholders show that leadership is shared between researchers and teachers, then transferred to students for most of the study (Fig. 4B). One of the most crucial impacts that transdisciplinary frameworks can deliver is the development of attitudes and agency in science and research^40,41,50^. By recognizing that assigning leadership roles to students is crucial, we foster long-term trust and confidence in research. Finally, by reintegrating our study process into the guiding transdisciplinary framework, we can evaluate efficacy and identify potential areas for improvement in designing and delivering impact for such initiatives (Fig. 4C). This helps evaluate action- and actor-oriented considerations for similar programs^51^. Collectively, these findings demonstrate that a strategically structured, flexible, and participatory stakeholder network can not only sustain engagement and knowledge exchange across multiple program phases but also fosters lasting scientific agency and capacity within the community.

**Figure 4.**
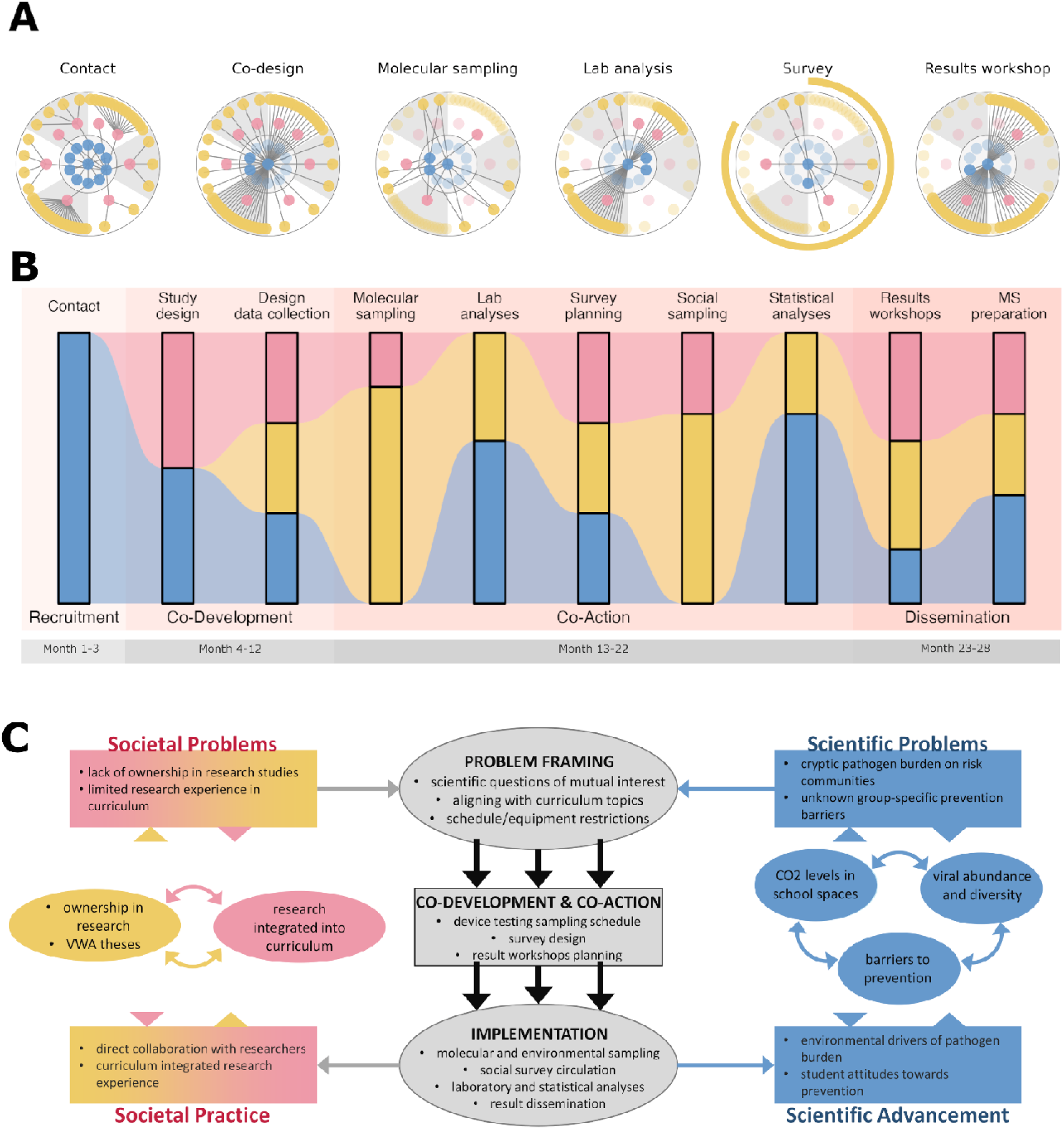
**Flexible network dynamics and distributed leadership support sustained engagement**. Network visualizations depict interactions (edges) among individual participants (nodes), with researchers in blue, teachers in pink, and students in yellow. Node positions are static to highlight changes in interactions over time, while inactive participants are faded at each stage. Edge dynamics reveal key stakeholders in recruitment (A – Network 1, edges represent initial interactions derived from email logs), demonstrate the importance of flexible engagement across stages (A – Networks 2–4, edges represent co-attended events based on shared calendars, and Discord discussion threads), illustrate the capacity of citizen science to amplify response rates (A – Network 5, edges derived from shared calendars), and show how outcomes can attract new participants to sustain continuity (A – Network 6, edges represent co-attendance from attendance sheets). Temporal shifts in engagement among stakeholder groups (B) confer leadership and agency to students within school-based citizen science programs. Mapping these processes (lowercase text) onto elements of the transdisciplinary framework (uppercase text) enables evaluation of the effectiveness of transdisciplinary implementation.

## Discussion

Recent studies show a critical role for co-developmental citizen science in public health and biomedical research in improving community health^52^, contributing to population data^53,54^, and interrupting health inequities^55^. By unifying environmental, molecular, and socio-epidemiological data into a single, harmonized framework, this study establishes a flexible and scalable model for transdisciplinary epidemiology that addresses critical gaps in traditional monitoring and informs integrated public health strategies.

Pathogen abundance measurements showed differences in availability and applicability across the different sample types. Air filter and swab samples are collected by involved students, but handkerchief samples require a large-scale collaborative effort from each user to deposit their disposed tissues into the designated container, leading to irregular sample availability. At the same time, swabs and air filters sample airborne pathogen concentrations, while disposable handkerchiefs are deposited by individuals who are largely symptomatic, assuming a higher rate of viral shedding and concentration^56,57^. Air filters and surface swabs are therefore more representative of airborne viral abundance than handkerchief samples, an essential result for replicating and/or upscaling our program, and selecting methods with the highest added value and efficiency.

Temporal analysis of air filter and surface swab samples revealed dynamic shifts in pathogen abundance, with RSV predominating in early-phase schools and SARS-CoV-2 in later phases, mirroring trends observed in regional hospital data. Across sample types, viral richness remained consistently high, demonstrating that diverse molecular sampling approaches can robustly capture the breadth of respiratory pathogens in school environments. The consistency of these school-based measurements with conventional molecular surveillance highlights their suitability for direct comparison and integration into broader public health monitoring frameworks. Since increasing CO_2_ levels were associated with increasing viral abundance but not richness, this suggests that ventilation has little effect in decreasing viral introduction into schools. However, once a pathogen has entered congregated school spaces, longer time spent in elevated CO_2_ levels will lead to increased transmission and an elevated risk of infection. Ventilation, a key measure in maintaining good indoor air quality, is therefore less likely to affect the number of pathogen species introduced into schools, but can influence the viability and transmissibility of those already circulating among students.

Survey data revealed that while students widely recognize vaccination and other preventive measures as effective, actual and perceived compliance—particularly for social distancing and absenteeism—remains limited by social and structural constraints. These results underscore the importance of identifying behavioral and contextual barriers to implementing prevention strategies in schools, offering actionable insights for designing more effective, student-centered public health interventions. When asked about how symptoms influence decisions regarding absenteeism, fever and sore throat were the decisive factors for staying home, not nasal discharge or coughing. Though unfortunately not unexpected, this result is in stark contrast to the viral shedding and infectivity, which are typically highest during coughing and nasal discharge^58^. Education of students and parents about transmission rates during different symptoms could help improve preventive efforts.

These results demonstrate the transformative potential of transdisciplinary frameworks for infectious disease management, in which bidirectional exchanges between researchers and communities generate high-quality data and potentially long-term societal impact. By linking scientific inquiry with community participation, such models create resilient knowledge networks that enhance preparedness, trust, and long-term capacity for public health response.

The LuPFiS program illustrates how community science can serve as a powerful approach to co-design epidemiological surveys with vulnerable communities, thereby advancing the emerging concept of transdisciplinary epidemiology. By integrating molecular and socio-epidemiological methods within a participatory framework, the program generates consistent, comparable data streams while simultaneously enhancing community engagement through assigned leadership roles. Unlike conventional surveillance systems, LuPFiS delivers environmental, molecular, and social data from parallel sampling—a combination essential for comprehensive infection monitoring that is rarely achieved in practice^59^.

Clinical datasets are often biased towards individuals with access to healthcare, wastewater signals are insufficiently resolved to capture group-level variation, and census or online data lack molecular resolution. By sourcing all three data types concurrently within high-risk communities (Fig. 5), transdisciplinary epidemiology fills a critical gap in current surveillance approaches (Supplementary Table 1).

**Figure 5.**
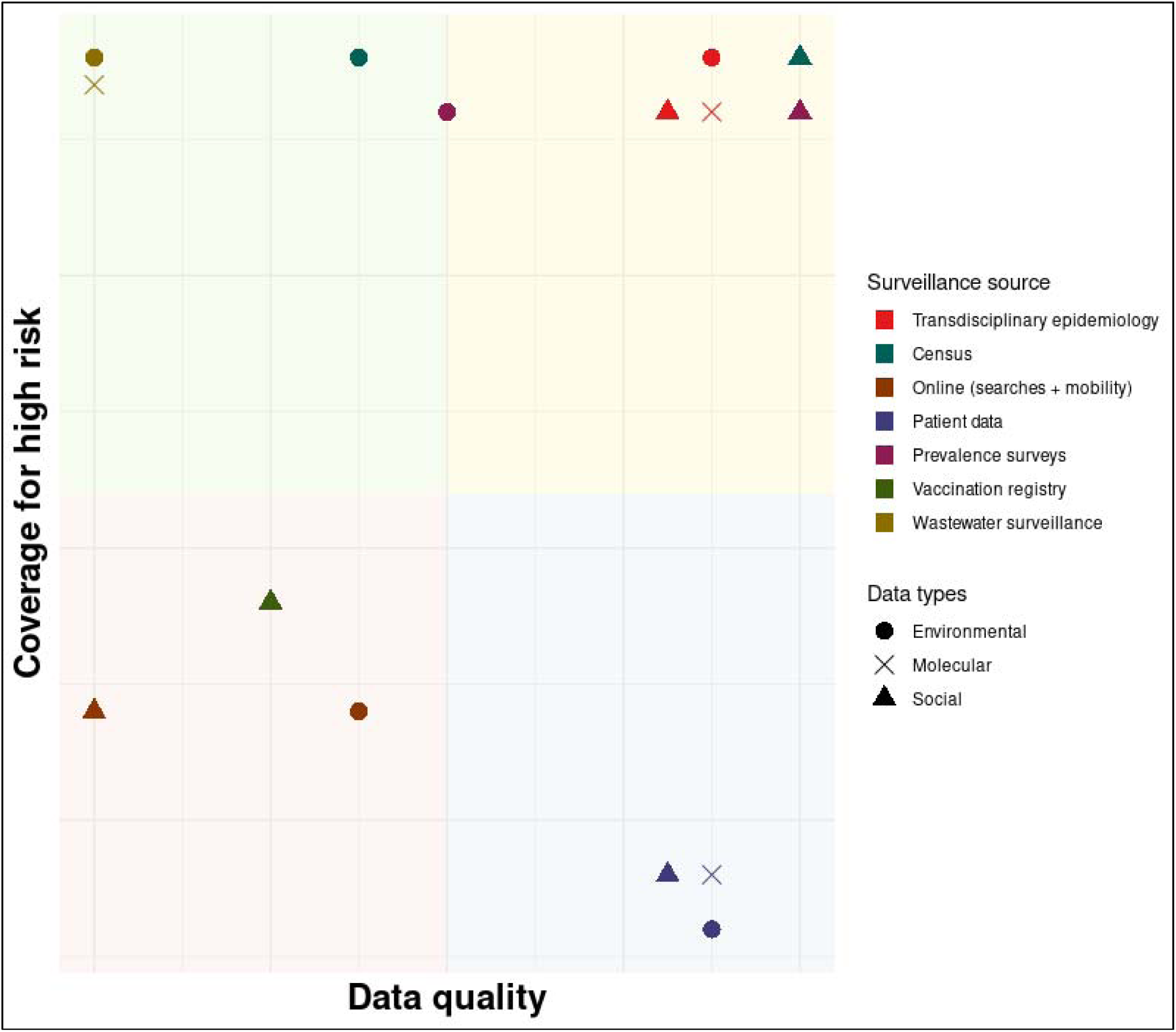
Transdisciplinary epidemiology delivers high quality, comprehensive epidemiological data for otherwise under surveyed risk groups. Data sources for epidemiological surveillance data (colour legend) are shown on an estimated scale for data quality and coverage for high risk populations. Each surveillance data source is represented by the data types (shape) it is able to collect. Sources delivering high quality data with high coverage for risk groups generally deliver social and environmental, but rarely molecular readouts (census, online search and mobility data). Patient data represent all data types in high quality, but is typically very low in coverage for risk groups, while wastewater surveillance delivers high coverage environmental and molecular data is bad quality. Finally, voluntary prevalence surveys as well as vaccination registries are restricted to social and limited environmental data of low coverage and quality. The only data source delivering all types in high quality and coverage is transdisciplinary epidemiology. Plot modified from Kraemer et al. 2025

The resulting data can be fully integrated with established surveillance databases, providing a compelling case for scaling school-based participatory science as a model for infectious disease monitoring. Molecular analyses demonstrated that community science in schools can deliver sustained pathogen data, with consistent detection of respiratory pathogens across participating sites. Sample robustness and reproducibility enabled seamless integration into molecular pipelines typically reserved for patient-based or wastewater sampling, underscoring the capacity of citizen-derived data to complement conventional surveillance systems and inform public health decision-making^38^. Considering uniform sampling efforts in each institution, these analyses can help distinguish schools facing a higher pathogen burden and therefore an elevated risk of infection^15^. Our study aligns with recent results pointing to air sampling and environmental surveillance being a scalable, high throughout surveillance tool for acquiring sequence data from congregate settings^15,60^. In parallel, social surveys capitalized on the community connections and network established during recruitment, yielding response rates more than five times higher than direct student participation in survey design.

An integrated community science approach, such as LuPFiS, offers high-resolution insights into barriers to policy implementation and provides an evidence base for targeted interventions. By assigning leadership roles to students, the program also fostered engagement and empowerment, echoing prior findings from participatory science initiatives^41,50,61^. Embedding outcomes within a transdisciplinary framework further enables iterative refinement of collaborative models, strengthening reciprocal exchange between science and society. The resulting integrated datasets align with the evidence needs of the UN Sustainable Development Goals, many of which require granular, community-rooted data to track progress effectively. By actively involving citizens in both data generation and interpretation, transdisciplinary epidemiology helps bridge persistent engagement gaps that have hindered global SDG advancement^35,38,53^. Together, these findings demonstrate that transdisciplinary epidemiology can deliver valuable complementary datasets that are applicable and scalable for inclusive, high-resolution epidemic preparedness.

## Methods

We recruited six public high schools from the Viennese metropolitan area (Supplementary Figure 5). 47 students (14-18 yrs) and eight teachers worked alongside researchers to co-design and conduct molecular and socio-epidemiological sampling and analysis. Study phases generated environmental, molecular, and social samples and readouts, with differential stakeholder engagement.

### Molecular epidemiological data collection

Sampling was conducted February-April, 2023, in a single room of six different high schools in the Viennese metropolitan area (Supplementary Figure 5). It was performed in two consecutive phases: Phase 1, involving Schools A-C over weeks 1-5, and Phase 2, involving Schools D-F over weeks 6-10. Sampling stations were set up in locations chosen together with students and teachers. Stations were placed in classrooms (4 schools), the cafeteria (1 school), or the break room (1 school). Each station included an air sampler, a bin for handkerchief samples, and a CO_2_ monitoring device (AIRCO2NTROL 5000, TFA Dostmann GmbH & Co. KG, Germany) that recorded CO_2_ levels every 5 minutes. Air samples were collected using the AerosolSense™ air sampler (Thermo Fisher Scientific, AEROSOLSENSE), a high-throughput device designed for continuous environmental aerosol monitoring^62^. Sampling was conducted continuously over 24 hours. Upon completion of sampling, cartridges were removed, sealed, and stored at room temperature until same-day pick-up. Participants also collected used disposable paper tissues in a designated bin and surface swabs from door handles of the sampling location using DNA/RNA Shield collection tubes (Zymo Research, R1107). All samples were then transported to the laboratory and extracted on the same day (see Supplementary material for process description). Molecular samples were collected exclusively by student and teachers, samplers were operated, cartridges exchanged, and metadata entered into the online database by teachers and students, with researchers providing remote support.

Viral abundance was quantified by digital PCR (dPCR) with multiplex panels targeting SARS-CoV-2, RSV, and influenza A and B, followed by hybrid-capture metagenomic sequencing of PCR-positive samples to assess viral richness (see Supplementary Methods for a detailed description of the analytical workflow).

### Environmental data collection

Concurrently with molecular sampling, we placed CO_2_-measuring devices (AirCo2ntrol 5000, TFA Dostmann, Germany) at all sampling stations throughout the sampling period in each school. CO_2_ concentration was recorded every 5 minutes and exported in parts per million (ppm) units. Data was saved on the built-in SD cards and extracted at the end of sampling in each school. CO_2_ data were subset to the 36-hour window preceding each metadata upload to align measurements with the timing of molecular sample collection.

To characterize COLJ levels, we quantified the Daily duration of high-risk exposure as the total time during which concentrations exceeded 1400 ppm^63,64^, a threshold associated with symptoms such as drowsiness and fatigue.

To further link pathogen sampling to air quality during the surrounding time period, we calculated 36-hour exposure to CO_2_ as the area under the CO_2_ curve measured within a 36-hour period preceding sample metadata upload. This enabled the estimation of net amount of CO_2_, correcting for the effects of long-term sub-threshold exposure. These variables tested impact of air quality on pathogen abundance and diversity for each school.

### Socio-epidemiological data collection

We used quantitative methods to investigate the community’s knowledge of transmission and to evaluate the feasibility of public health measures and awareness of infection risks. We co-developed an online survey with student citizen scientists to assess students’ perceptions and attitudes towards prevention measures. Data mapped their perceptions of efficacy, compliance, feasibility and peer-compliance with vaccination, social distancing, absence during sickness, public health announcements in the media, and indoor ventilation as school-based prevention measures. We further asked students to determine which of their social circles has the greatest effect on their decision to vaccinate. Questions used a Likert scale from 1-5, with 1 indicating total disagreement and 5 indicating total agreement with the statements.

Surveys were built on the SoSci Survey (SoSci Survey GmbH, Germany) platform and distributed from September – November, 2023 to all students of participating schools via QR codes. Surveys were fully anonymous and age-appropriate, thereby subject to an ethics waiver (Medical University of Vienna, 1219/2024). Data from the platform was downloaded to the virtual servers of the Medical University of Vienna, in accordance with the highest standards of data security.

### Transdisciplinary process

We developed and applied the three steps workflow RECRUITMENT-DEVELOPMENT-ACTION to involve teachers and students throughout the entire research process. RECRUITMENT was initiated through non-selective invitations sent to approximately 20 Viennese high school teachers in the network of Open Science – Life Sciences in Dialogue and the Vienna Open Lab, an Austrian science education institution. Enrollment in the program was completely voluntary. Teachers were invited to one of two 90-minute info sessions introducing program goals and the research team.

Those who had signed up at the end of the info sessions were then enrolled into the program and progressed to the CO-DEVELOPMENT phase. The first part of co-development focused on aligning scientific interests with schools educational priorities to formulate final research questions, as well as benchmarking and designing the study timeline and methodology. Furthermore, together with teachers we explored ways to integrate the program into the curriculum to foster engagement and impact among students. Following their guidance, we developed options for both types of mandatory research projects in the Austrian high school curriculum: the Pre-academic thesis (“Vorwissenschaftliche Arbeit”; VWA) is an in-depth research essay, while the Final work (“Abschließende Arbeit”; ABA) allows the use of different media formats (videos, podcasts, etc.) to summarize a project. Students enrolled in our program could choose to write their essay based on the work conducted in our program, with support from the research team. In the second part of the co-development phase, teachers recruited students from their schools, who joined the process of designing a data collection strategy for both molecular and socio-epidemiological methods. We held workshops for all enrolled students, providing training in both types of methods, followed by a discussion about study design. During the CO-ACTION phase, molecular data collection was tailored to both local conditions and school schedule, devices and collection stations were deployed in jointly determined school spaces, and timing of sampling was adjusted to individual student schedules. Air filter devices were benchmarked to ensure consistent detection across sample types (Supplementary Figure 6). Socio-epidemiological surveys were co-developed with students and teachers, using their insights to formulate questions and response options. Students were introduced to laboratory analyses through two workshops, where they conducted extractions on mock samples and explored PCR and sequencing workflows. Those pursuing their VWA within the project framework later contributed to statistical analyses of the dataset. Finally, during the DISSEMINATION phase, we fed results back to school communities through on-site workshops and Q&As, and developed the manuscript with teachers’ and students’ feedback.

To facilitate communication between the research team and citizen scientists, we used Discord, a platform familiar to a large proportion of students. This proved highly efficient for real-time troubleshooting, scheduling sampling dates, discussing emerging issues, and sharing materials and experiences. We also used digital data such as Discord project site traffic, together with meeting logs to quantify, track and analyze stakeholder engagement dynamics throughout the study period.

### Data Analyses

We examined how viral abundance varied across schools and sample types to identify potential environmental and temporal drivers of pathogen dynamics. Relative pathogen abundances were derived from digital PCR data and compared within and between schools, as well as to hospitalization rates from Vienna within the sampling weeks^44^. Exploratory analyses, including non-metric multidimensional scaling and permutation tests, were used to assess the influence of school, sample type, and collection time on viral abundance patterns. To account for zero-inflated and skewed data distributions, we applied generalized linear models of the Tweedie family^65^ to separately test for detection probability and abundance intensity. Full details of the statistical procedures and model specifications are provided in the Supplementary Methods.

Sequence data determined the diversity and distribution of respiratory viruses detected in school environments. Viral richness, defined as the number of distinct species identified per sample, was used as the primary diversity metric. We first explored the overall structure in the data using principal component analysis (PCA). Subsequently, we tested the influence of school and sample type on viral richness with generalized linear models. Post-hoc comparisons were performed to resolve pairwise differences while controlling for school effects. Detailed model specifications and statistical procedures are provided in the Supplementary Methods.

Indicators of indoor air quality, expressed as duration of daily high-risk CO_2_ levels and cumulative 36-hour CO_2_ exposure, were associated with viral abundance and diversity in schools. Since the two CO_2_ metrics were positively correlated, they were analyzed in separate models to avoid collinearity. Environmental pathogen abundance (air filter and swab samples) was modeled using generalized linear models with a Gamma distribution and log link, while viral richness was analyzed using Poisson models. School was included as a covariate in all models. Comprehensive descriptions of model structures, assumptions, and diagnostics are provided in the Supplementary Methods.

Survey responses were analyzed to assess students’ perceptions and behaviors regarding infection prevention measures. Individual responses on Likert scales were aggregated into four variables representing perceived efficiency, personal compliance, feasibility, and perceived peer compliance across five prevention domains (vaccination, social distancing, absence during illness, public health messaging, and indoor ventilation). Differences among these variables were tested using repeated-measures ANCOVA, with respondent sex included as a covariate. Detailed statistical procedures and model diagnostics are described in the Supplementary Methods.

To gain a better overview of the transdisciplinary process integrating research directions, methods, and stakeholder interests, we employed network visualization to track recruitment patterns and engagement across study phases. Data extracted from attendance records, shared calendars, and Discord discussion threads were used to quantify the timing and frequency of individual contributions in joint events and online interactions. These metrics enabled estimation of stakeholder involvement across study components as a metric of ownership for each stakeholder group. Finally, we mapped individual design elements of the study back onto the transdisciplinary framework to evaluate implementation and design future guidelines for repeatability and upscaling.

During the preparation of this work the author(s) used ChatGPT (OpenAI, GPT-4) in order to streamline wording, shorten sentences, and fine-tune paragraph structure for clarity and conciseness. After using this tool/service, the author(s) reviewed and edited the content as needed and take(s) full responsibility for the content of the published article.

### Limitations

We note that analyses may be underpowered due to low number of schools involved. Extending sampling periods or repeating sampling across different seasons will increase power to compare school data with wastewater and hospital-derived datasets and to reveal seasonality within the same school. Increasing the number of participating schools within a region provides robust data on the infection risks faced by the adolescent community, while covering larger geographical areas allows comparisons across urban and rural schools. Our pilot study recruited schools voluntarily, implying that participating schools had already had a favorable attitude towards research engagement. A future direction of development is to capitalize on teachers’ knowledge and experience in the LuPFiS program and to design a framework for schools serving low-income communities. Finally, increasing capacity would allow for establishing multiple sampling stations within schools to detect effects linked to different uses of indoor spaces.

## Data Availability

The datasets generated and analyzed during the current study include environmental measurements, molecular pathogen data, and survey responses collected in school settings, as well as contact information of participants for tracking interaction networks and school-related metadata. De-identified and anonymized environmental, molecular and social datasets are available upon request. Anonymized subsets of interaction data as well as school metadata can be made available upon request, subject to considerations of maintaining full anonymity for schools throughout the analyses. Personal data of participants is not available due to ethical and privacy restrictions involving minor participants but an anonymized subset may be made available from the corresponding author upon reasonable request and with appropriate approvals.

## Acknowledgements

We thank all students, teachers, and staff who assisted us in designing and implementing our programs in their communities. We are also thankful to Prof. Regina Sommers for her help with laboratory analyses during the device benchmarking tests. All activities conducted within our program are subject to an ethics waiver issued by the Ethics Committee of the Medical University of Vienna (EC#: 1219/2024). This study was funded by grants from the Austrian Science Fund (Top Citizen Science initiative #TCS153 and Cluster of Excellence #COE7 to AB).

## Author contribution statement

O.M. and A.B. conceptualized the study. O.M., I.G., B.G. and K.G. designed the transdisciplinary methodology. Study design was carried out by O.M., A.S., D.G., B.G., K.G., S.P., B.G, V.K., B.S., and D.H. D.G., A.G., F.S., N.B., carried out data collection, including environmental sampling, molecular analyses, and survey implementation, with the supervision of T.F. and O.M. O.M., A.S., T.F., M.T., F.A., I.G. and A.B. performed data curation and formal analysis, assisted by H.R., A.S., M.T., J.S., V.D., W.O. and C.Ma. O.M. wrote the original draft. All authors contributed to interpretation of the data and critical revision of the manuscript. O.M. and A.B. supervised the project. A.B. secured funding.

## Supplementary material

### Sample preparation and Nucleic acid extractions

Using sterile tweezers, collection substrate from the air sampler cartridges were transferred and washed in 1:1 1 mL Phosphate-Buffered Saline (PBS) and DNA/RNA Shield (Zymo Research, R1200-25) solution. The collection substrate was pushed down using the sterile tweezer or a pipette tip to ensure even distribution of liquid, followed by vortexing for 10 seconds. 900 µl of resulting suspension was aliquoted into three tubes (300 µl each). Carrier RNA (5.6 µL of a 1 µg/µL solution) was added to two of the triplicates to enhance nucleic acid recovery, particularly for downstream qPCR applications. The third triplicate, intended for sequencing, was processed without carrier RNA to avoid contamination of sequencing libraries. Nucleic acid extraction was performed using the manufacturer’s protocol for the Quick-DNA/RNA Viral Kit (Zymo Research, D7021), with the exception that beta-mercaptoethanol was deliberately omitted to reduce exposure to toxic reagents and improve laboratory safety. Final elution was carried out in 50 µL of nuclease-free water.

For paper tissue samples, tissues were first inspected for moist residues. If present, swabbing was performed with DNA/RNA Shield–drenched swabs, and the swab liquid was retained for later pooling. Each tissue was placed in a laboratory plastic waste bag and incubated with 6 mL of DNA/RNA Shield (or 10 mL for single tissues) in a Stomacher device at room temperature for at least 10 minutes until foam was observed. The homogenate was filtered through a RNAse/DNAse-free ceramic filter into a 50 mL falcon tube. If residual debris remained, filtration was repeated with an additional 0.45 µm polypropylene membrane filter (Whatman® PPHH09050). If swabs were used, their fluid was added to the final filtered sample without re-filtration. The clarified sample was vortexed briefly, and 2.4 mL was aliquoted into three tubes (800 µL each). To each, 8 µL of Proteinase K [20 mg/mL] (Zymo Research, D3001-2-125) was added, followed by incubation at room temperature for 15 minutes. Samples were then purified using the same nucleic acid isolation kit and protocol as above.

Surface swabs collected in DNA/RNA Shield collection tubes (Zymo Research, R1107) were processed by removing 900 µL of the primary sample and aliquoting into three 300 µL fractions. These were then subjected to nucleic acid isolation using the same kit and conditions described above.

Sample preparation and extraction was conducted by the research team, with students invited to one of two, 3 hour long practical workshops, where they received training in extraction and interpreting PCR results, and then conducted the entire extraction process on samples spiked with unknown amount of SARS-CoV-2 naked RNA. Extracts were transported back to the lab and resulting PCR plots of quantification analyses were fed back to each school group for interpretation.

### Pre-screening by digital PCR

Pre-screening of collected samples was performed using the QIAcuity ® One Digital PCR System (Qiagen) in combination with two Qiagen multiplex Microbial DNA Detection Assay panels. Panel 1 included targets for Rhinovirus (DMA00378-F), Respiratory Syncytial Virus – RSV (DMA00379-H), Adenovirus (DMA00371-R), and Influenza A (DMA00373-T) and B (DMA00374-C) virus. Panel 2 targeted Enterovirus (DMA00487-R), Norovirus [GII] (DMA00473-F), Human Coronavirus SARS-CoV-2 [N1] (DMA00710-C), Streptococcus pneumoniae (DMA00318-T), and Staphylococcus aureus (DMA00302-H). Reactions were set up following the QIAcuity ® OneStep Advanced Probe Kit (Qiagen, 250131) Quick-Start Protocol for multiplex detection (up to five targets) using 26K 24-well nanoplates (Qiagen, 250001) with a 20 µL RNA template input per reaction. All procedures were carried out according to the manufacturer’s instructions. A Microbial DNA Positive Control (Qiagen, 338135) was included in each run to validate assay performance.

### cDNA Synthesis, Library Preparation, and Sequencing

Complementary DNA (cDNA) synthesis was performed using the ProtoScript® II First Strand cDNA Synthesis Kit (New England Biolabs [NEB], E6560L) and the NEBNext® Ultra™ II Non-Directional Second Strand Synthesis Kit (NEB, E6111S). For first-strand synthesis, 5 µL [60 µM] of Random Primer 6 was combined with 15 µL of RNA sample, incubated at 65°C for 5 minutes, and immediately placed on ice. The first-strand synthesis reaction mix was prepared according to the manufacturer’s instructions, and 30 µL of this mix was added to each sample. The reaction was incubated at 25°C for 5 minutes, 42°C for 1 hour, and then 80°C for 5 minutes. Second-strand synthesis was then carried out using the same kit, following the manufacturer’s protocol. Samples were incubated at 16°C for 1 hour with open tube lids to allow evaporation as part of the protocol.

Following synthesis, double-stranded cDNA was purified using SPRI bead cleanup. Specifically, 80 µL of SPRI beads (Beckmann Coulter, B23317) were added to each 80 µL reaction, incubated at room temperature for 5 minutes, and washed twice with 200 µL of 80% ethanol. The purified cDNA was eluted in 28 µL of nuclease-free water. DNA quantification was performed using 1 µL of eluate and the Qubit™ High Sensitivity dsDNA Assay Kit (Thermo Fisher Scientific, Q33231), according to the manufacturer’s instructions.

Library preparation was conducted using the NEBNext® Ultra™ II FS DNA Library Prep Kit for Illumina (NEB, E7805L), following the manufacturer’s instructions. Based on cDNA input quantification, 11 cycles of indexing PCR were used for samples with input below 100 ng, and 6 cycles for all other samples. Final library clean-up was carried out using 0.9× SPRI beads. A total of 400 ng of each library was pooled for hybrid capture enrichment.

To target respiratory viruses, the Twist Bioscience Respiratory Virus Panel (Twist Bioscience, 103067) was used for hybrid capture, following the version 1 protocol of the Twist Hybridization and Wash Kit. Final enriched libraries were sequenced on an Illumina® NovaSeq 6000 System in paired-end mode with 100 bp read length. Viral detections were obtained using a mapping-based analysis pipeline with a set of filters to reduce unreliable detection calls.

### Statistical models for viral abundance

PCR data was used to calculate total pathogen abundance across schools, to visually compare trends to those of wastewater monitoring within the sampling period. We also calculated relative pathogen abundance for each sampling event of each school to compare within and between school trends.

We then conducted exploratory analyses on the abundance data to discover any potential effects of school, sample type of time of collection. We searched for observable alignments in the data and then overlayed different metadata to guide further significance analyses. As abundance data was heavily zero-inflated, we used Non-Metric Multidimensional Scaling (NMDS), a widely utilized approach to dealing with similar abundance data^66,67^. We ran analyses with both Bray-Curtis and Jaccard distances, and forced 200 iterations. We used to stress levels to determine the better fit. Although both were close to zero (0·000000002592 and 0·000125, respectively), Jaccard-distances returned a substantially higher value, so we used that fit in future analyses. Finally, we used an *adonis* test to see if school or sample type had an effect on our distance matrix. As sample type returned a significant effect on the distance matrix (R2 = 0·016, p = 0·002), we proceeded with building models describing the relationship between pathogen abundance and sample type.

To test for the effect on sample type, we first investigated the distribution of pathogen abundance as the response variable. Heavy zero-inflation excluded the use of Gaussian models, but the continuous variable was also unsuited to Poisson model family. Finally, extreme values made the data right-skewed by inflating means, so Gamma models were also inadequate to capture variance. We therefore decided to apply a Hurdle model, where we conducted separate analyses to test for effects on detection and abundance. We assigned a new binomial variable (Detection) to cases of not-detected (0; Pathogen abundance = 0) and cases of detection (1; Pathogen abundance > 0). We then used Generalized Linear Models (GLZ) with binomial function to run logistic regression, with Detection entered as response, and sample type as explanatory variable. To test effect of sample type on abundance, we subseted our data frame for cases with Pathogen abundance > 0 and used abundance in future models. Investigation of the subseted abundance data showed it was also heavily-skewed with few extremes, we therefore applied GLZ model of the Tweedie family, which allows any power variance function to improve fit and is fit for analyzing viral abundance data^68^. We entered Pathogen abundance as response and sample type as explanatory variable, with power of variance set to 1.5 corresponding to compound Poisson, non-negative with mass at zero type distribution and a log link function.

### Statistical models for viral richness

We used sequence data to identify viral species collected from school spaces. By comparing count numbers for positive identification of each target, we determined the frequency of occurrence for each of the 24 identified viral species.

We characterized diversity with viral richness^69,70^, and conducted Principal Components Analysis (PCA) to search for axes or cluster representing underlying effect of metadata. Analyses returned 24 PCs with the first 9 explaining 73.15% or variance. PCA plots with overlayed metadata did not reveal any trend to be followed up on, we therefore conducted direct tests for analyzing any potential influence of schools and sample type.

Investigating the distribution of Richness variable revealed a Poisson distribution, we therefor built GLZ from Poisson family, with richness entered as response variable and Sample Type and School as explanatory variables. Both explanatory variables returned significant effects within the model, which were followed up by Tukey-tests for Posthoc analysis. Sample type no longer had an effect when data was corrected for School, but in line with expectations, School showed a significant result, we therefore entered School into all future analyses to correct for the effect.

### Statistical models for effect of CO_2_ levels

Effects of both Duration of daily high risk of CO_2_ levels and 36-hour exposure to CO_2_ were both tested in GLZ. We tested for correlation between the two variables using spearman correlation, and found significant positive correlation, we therefore analyzed effects in separate models to avoid collinearity.

Effects on pathogen abundance were investigated using GLZ from the Gamma family with log function. To correct for the significant effect of sample type, we subseted pathogen abundance data to exclude samples from handkerchiefs, and used only air filter and swab samples to create Environmental pathogen abundance variable. This was entered as a response into, along with either Duration of daily high risk of CO_2_ levels (Model 1) or 36-hour exposure to CO_2_ (Model 2), and school as a covariant.

Effects on pathogen diversity were also analyzed using GLZ from Poisson family with log function, and separate models were built for Duration of daily high risk of CO_2_ levels (Model 1) and 36-hour exposure to CO_2_ (Model 2), with school as a covariant. Since sample type had no effect on richness, data from all sample types was entered into the analyses.

### Statistical models for socio-epidemiological analyses

Social survey data was extracted to characterize individual preferences on the Likert scale for each question. Variables for each respondent characterized perceived efficiency (Efficiency), adherence to measures (Compliance), access to and ability to adhere (Feasibility) and perceived compliance among peers (Peer compliance) for each of the five prevention measures (Vaccination, Social distancing, Absence during sickness, Public Health announcements, Indoor ventillation).

Difference between the above-described variables was tested using Repeated Measures ANCOVA, following a normality test all involved variables. Gender of respondents was entered as a covariate to correct and test for effect. We conducted separate ANCOVa tests for all 4 prevention measures.

---

**Supplementary Figure 1.**
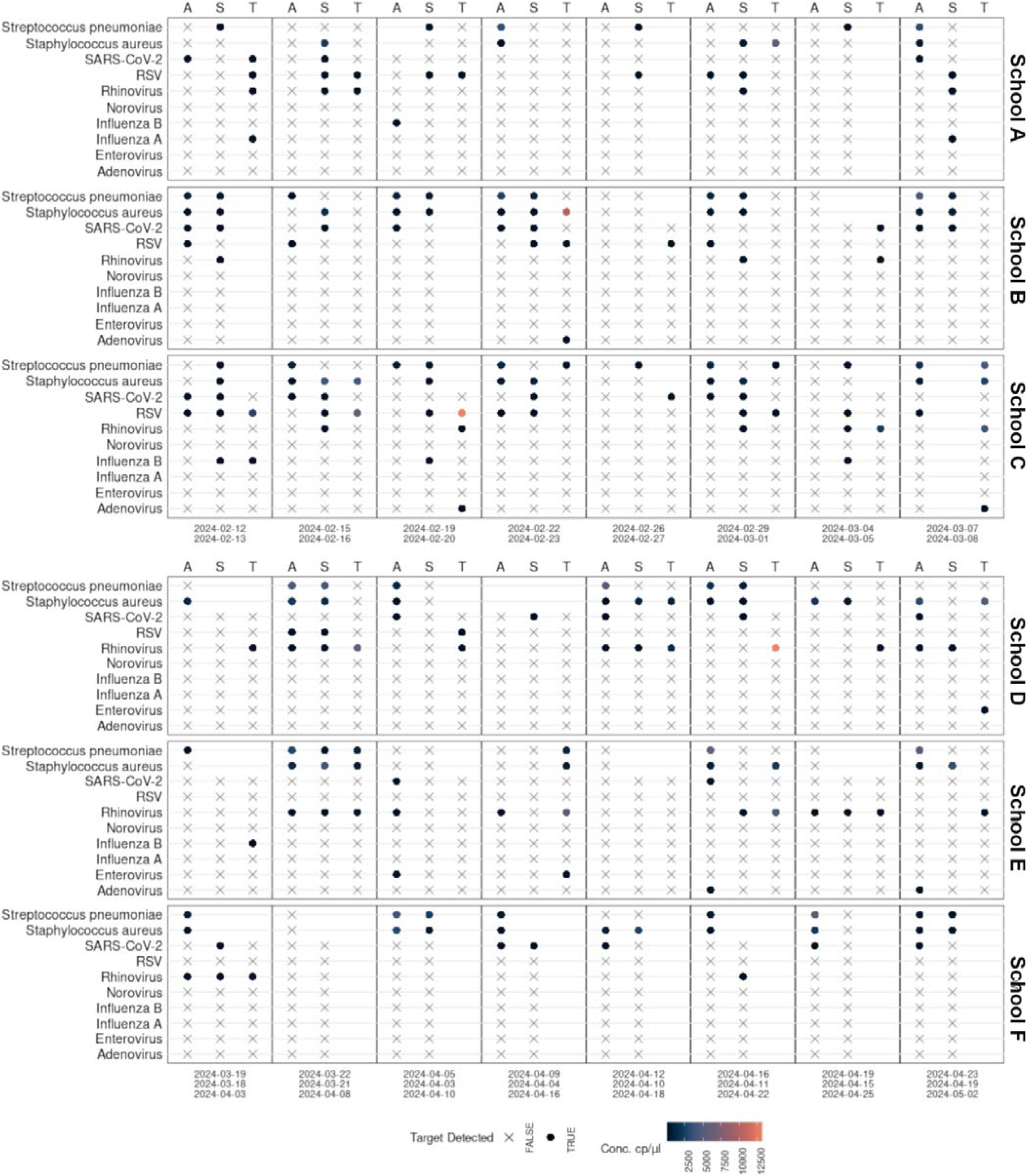
Detection of respiratory pathogens across six schools. Targets were measured over eight sampling events per school (A–F). Sample types on upper x axes are indicated by A (air filter), S (surface swab) and T (tissue sample). Each row corresponds to a pathogen target, with detection shown as filled circles and non-detection as crosses. Circle fill colour represents concentration (cp/µl) on a blue–red gradient. Dates of sample collection are shown on the lower x-axis.

**Supplementary Figure 2.**
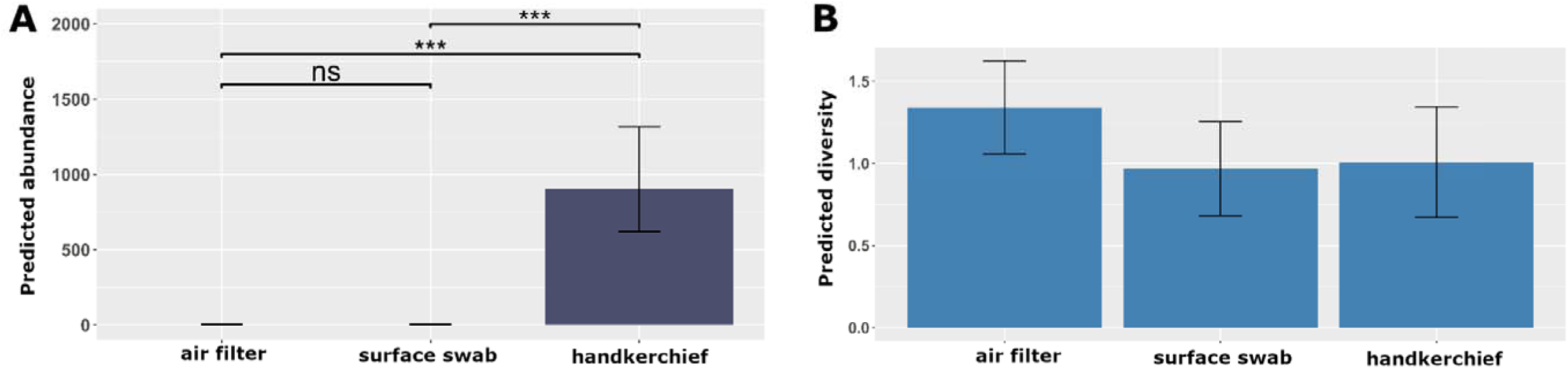
Effect of sample type on pathogen abundance and diversity. Results of generalized linear models testing the influence of sample type on predicted pathogen abundance (A) and diversity (B). Both response variables are represented by model residuals. Bars indicate mean estimates with standard errors. Significance levels are shown above comparisons (ns: not significant; ***: P < 0.001). Response variables are shown using model residuals.

**Supplementary Figure 3.**
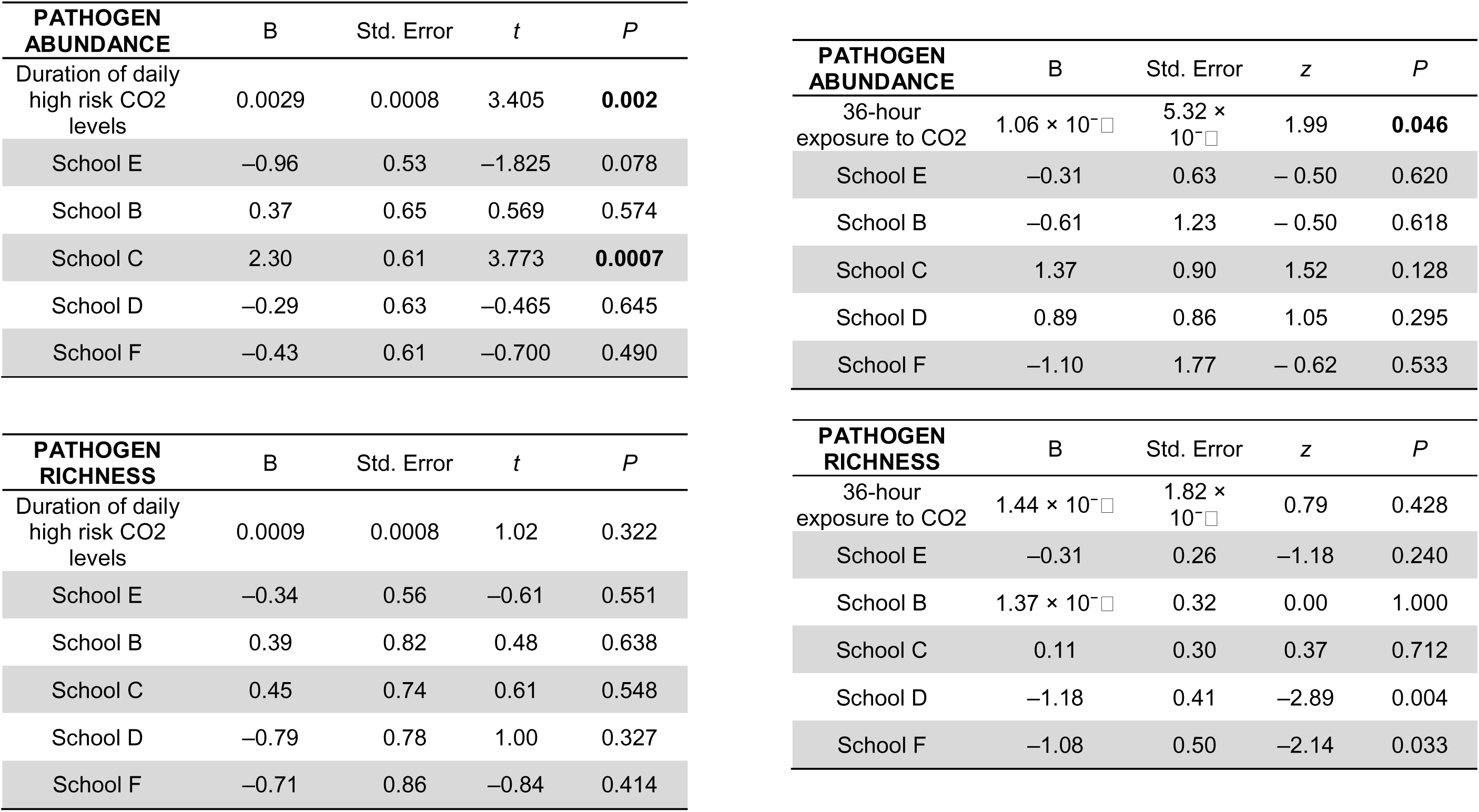
**Final models examining associations between CO**D **levels and pathogen abundance and richness.** Generalised linear models with a Poisson distribution and log link were used to estimate the effects of COLJ exposure metrics on pathogen abundance and richness. Models included school as a categorical covariate. Coefficients (B), standard errors, test statistics (t for duration-of-high-risk COLJ models; z for 36-h COLJ exposure models), and p values are reported. Significant associations (p<0·05) are shown in bold.

**Supplementary Figure 4.**
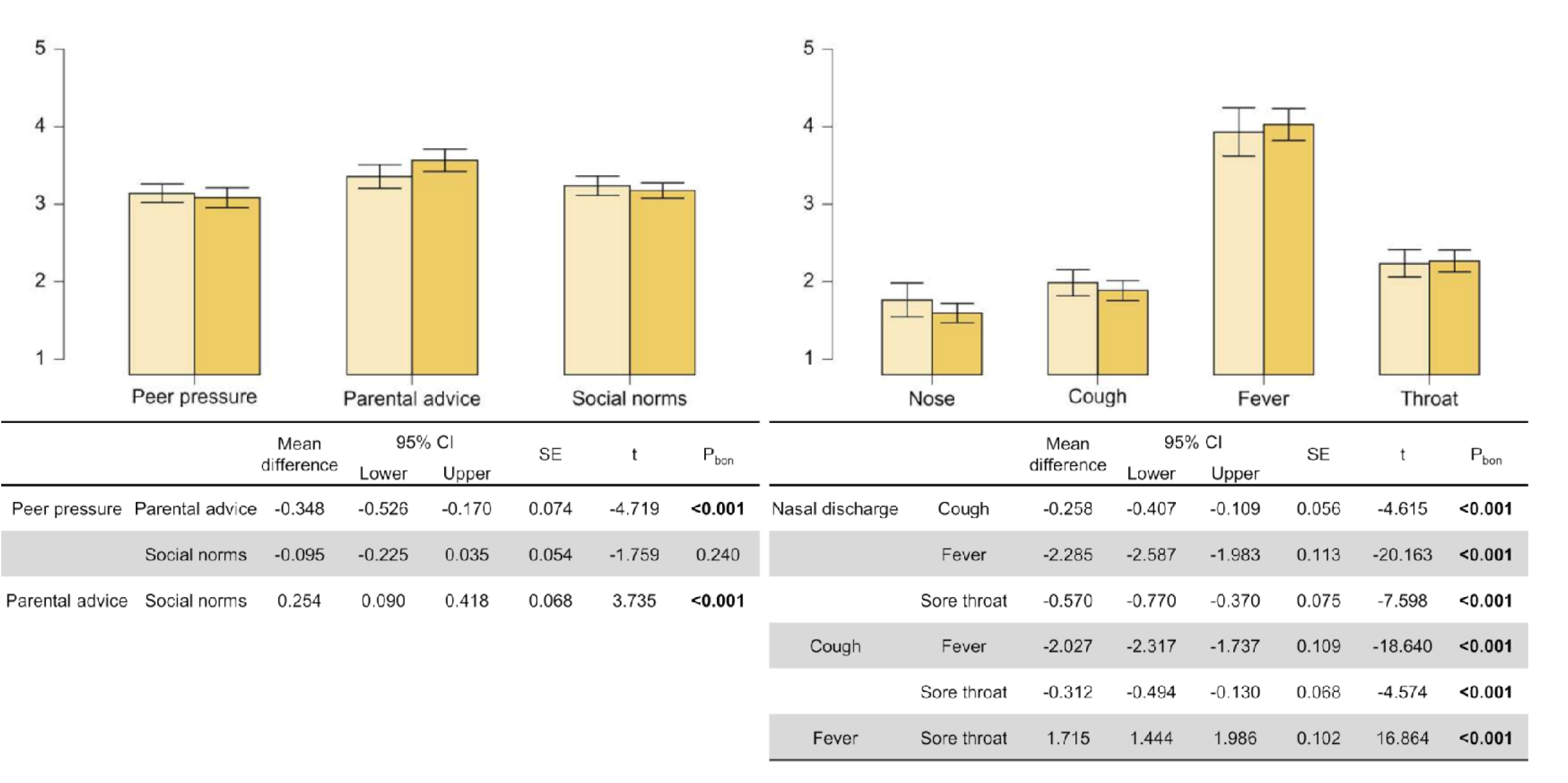
Factors driving vaccination decision and absenteeism. Results of generalized linear models testing the influence of social groups on vaccination decisions (A) and symptoms contributing to absenteeism (B). Both models represent response variables by model residuals. Bars indicate mean estimates with standard errors for Likert scales, effects are displayed for females (dark grey bars) and males (light grey bars) separately. Post-hoc tables display significance levels with Bonferroni corrections in bold (P < 0.05)

**Supplementary Figure 5.**
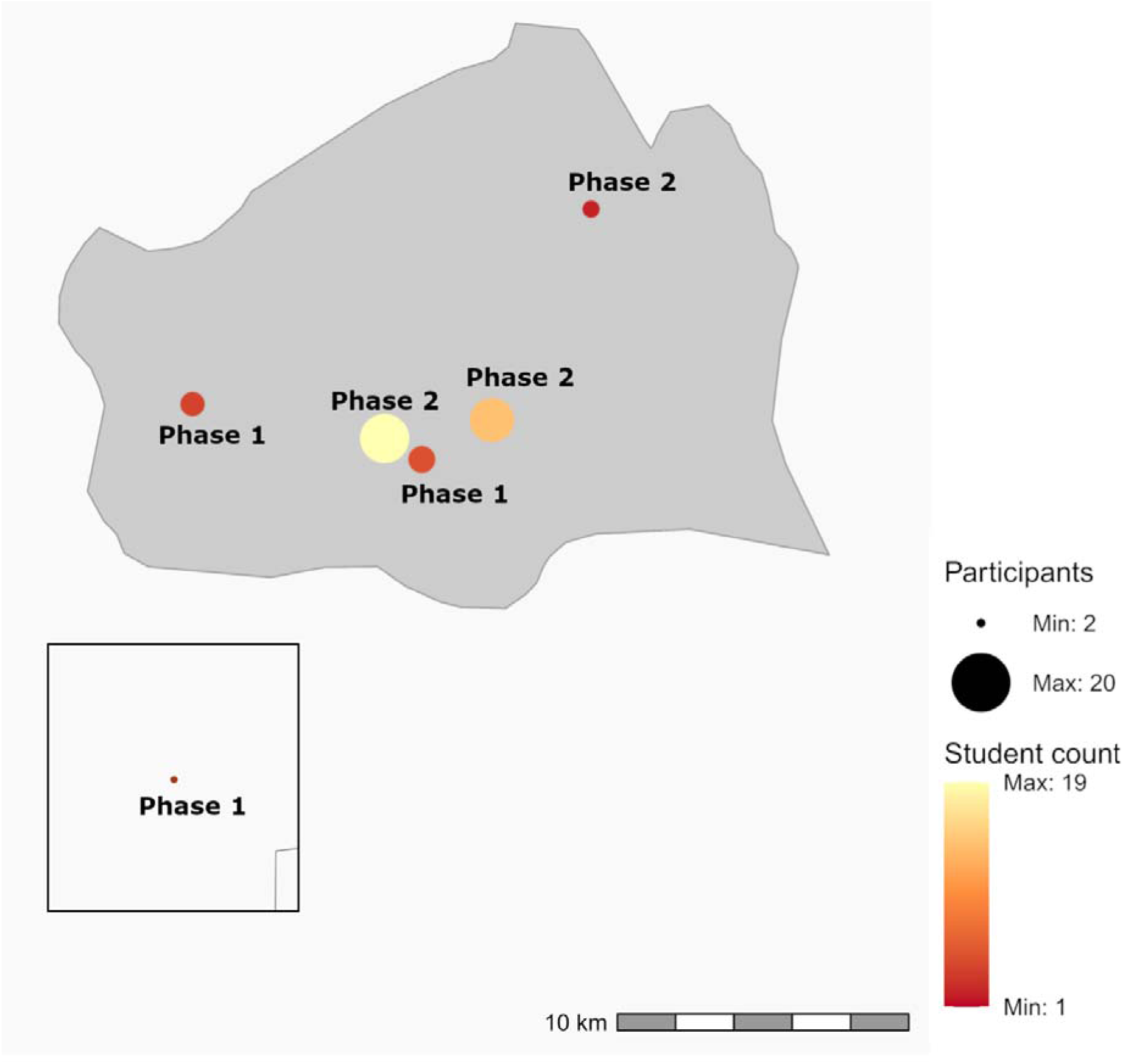
Schools enrolled in the LuPFiS program in the Viennese metropolitan area. Enrolled schools are shown on the map of Vienna (centre) and Wiener Neustadt (inset), with the sampling phase indicated next to each point. Point size reflects the total number of participants (students and teachers) at each school, and the yellow–red colour scale denotes the student–teacher ratio.

**Supplementary Figure 6.**
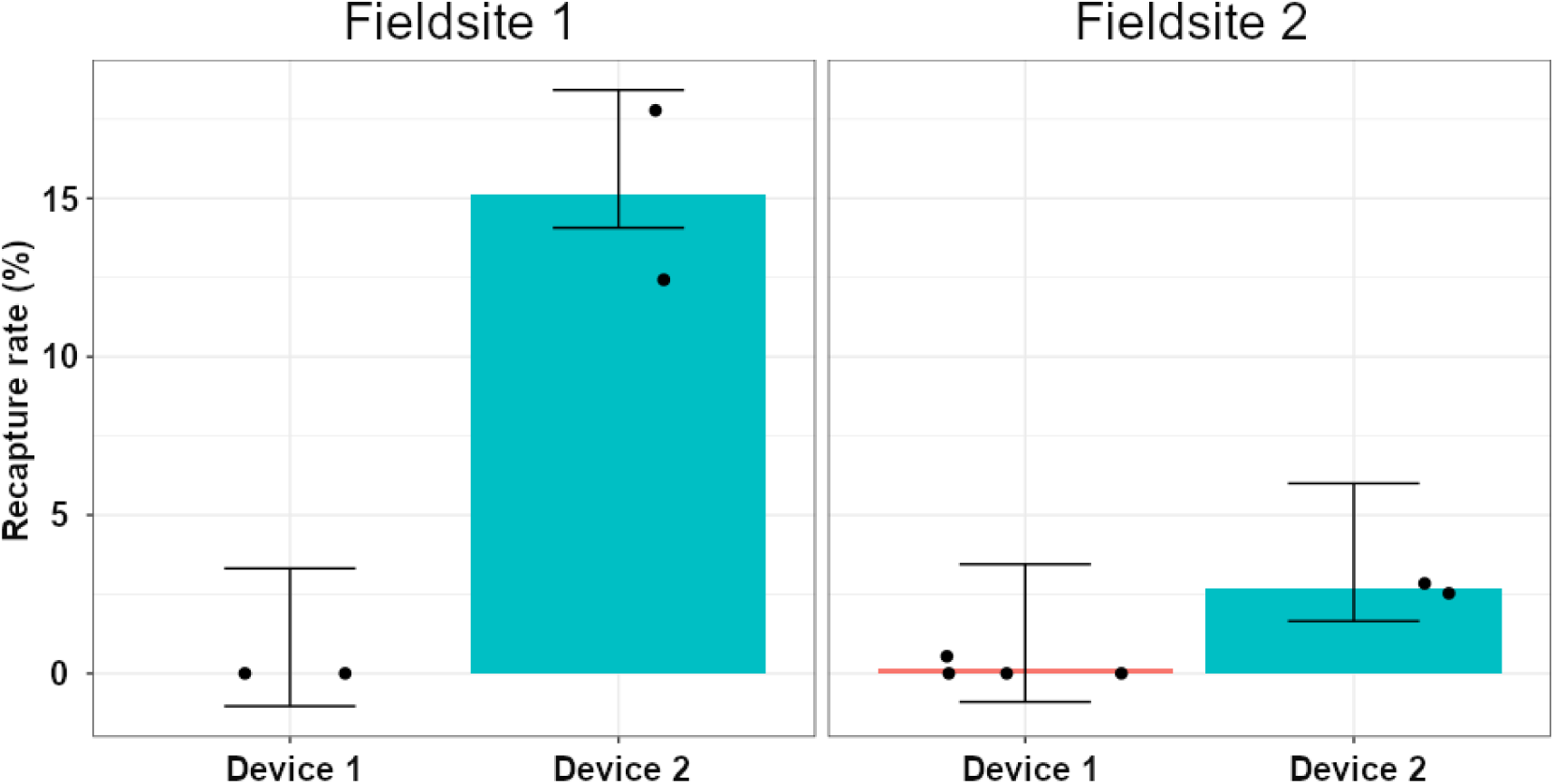
Benchmarking of air filter devices. Recapture rates of PhiX DNA were compared between two air filter devices operated for 24 h at two field sites following aerosolization of 1.08 × 10ℒ PhiX DNA copies in PBS buffer. Cartridges were extracted and analyzed by dPCR, with recapture rates calculated as detected copies divided by total input. Field tests showed higher recovery for Device 2 (AerosolSense, Thermo Fisher). Bars show mean recapture rate, whiskers show errors, data points are off center to avoid overlaps.

**Supplementary Table 1.**
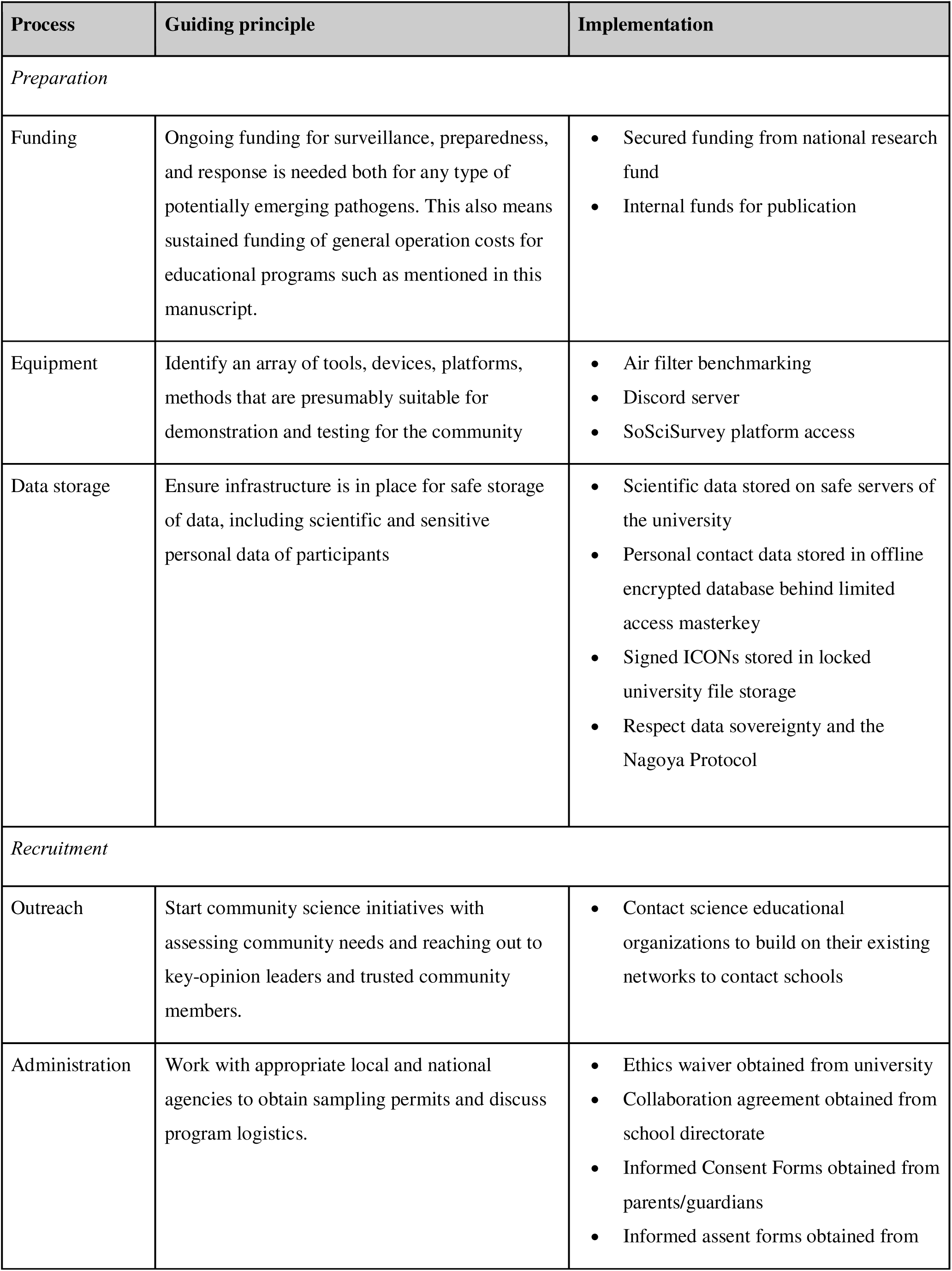
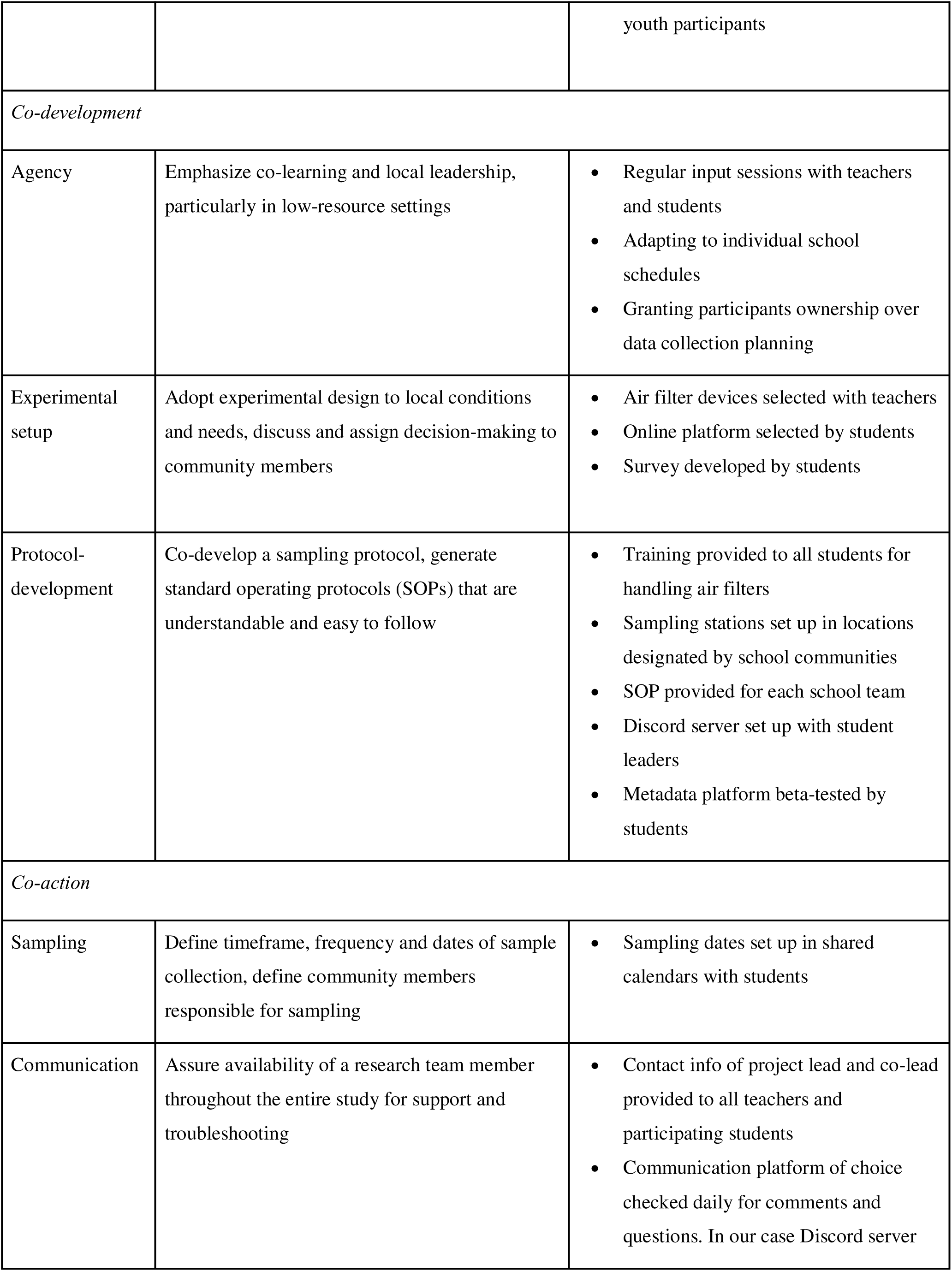
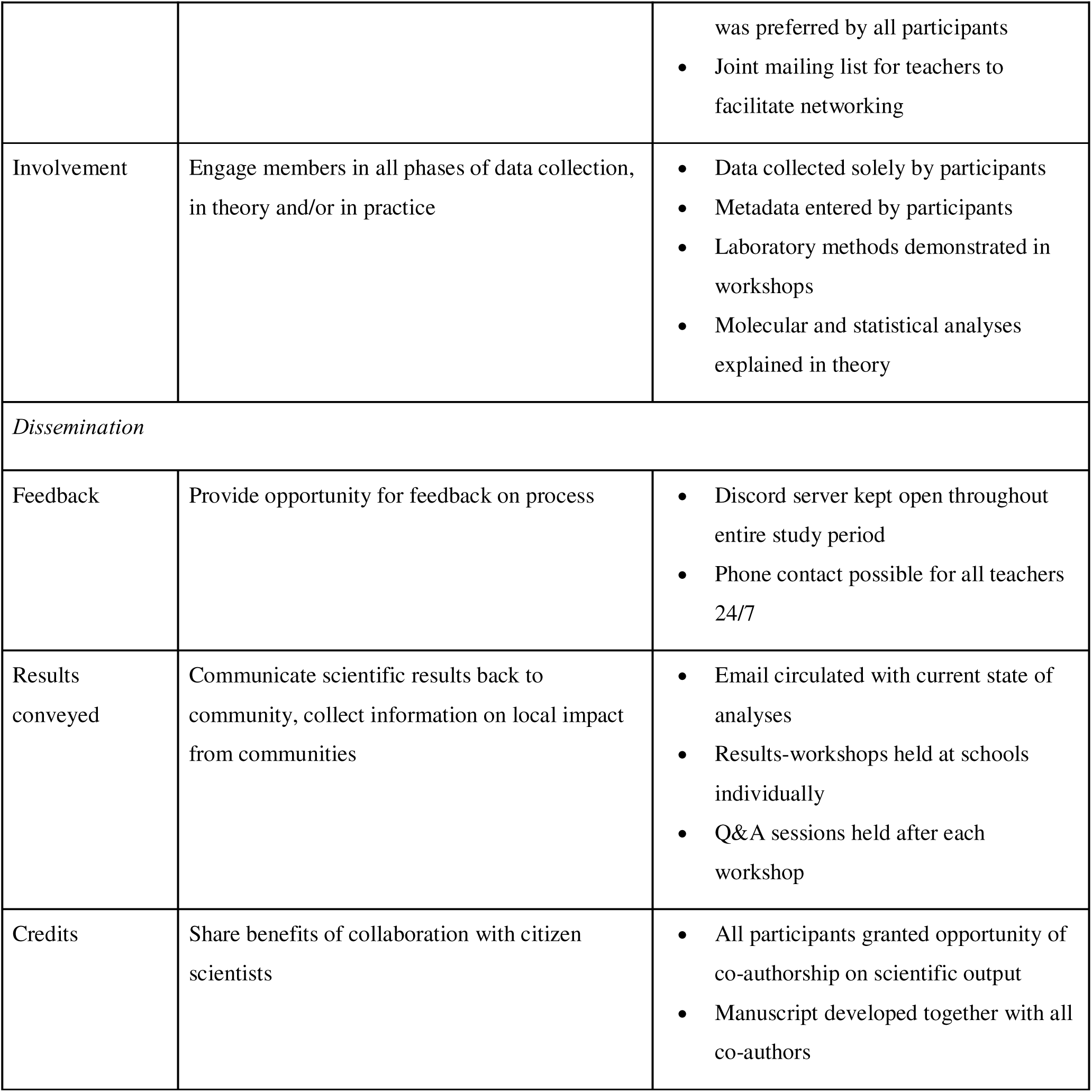
Overview of processes, guiding principles, and implementation steps establishing programs in transdisciplinary epidemiology. The table summarizes the main operational components of the study—preparation, recruitment, co-development, co-action and dissemination phases—and outlines how guiding principles were translated into practice. Implementation examples include approaches to funding, equipment setup, data management, participant engagement, and experimental protocol development.

